# Estimated Seroprevalence of SARS-CoV-2 Antibodies Among Adults in Orange County, California

**DOI:** 10.1101/2020.10.07.20208660

**Authors:** Tim A. Bruckner, Daniel M. Parker, Scott M. Bartell, Veronica M. Vieira, Saahir Khan, Andrew Noymer, Emily Drum, Bruce Albala, Matthew Zahn, Bernadette Boden-Albala

**Author notes:** **Corresponding Author:** Tim Bruckner, PhD, Associate Professor of Public Health, 653 E. Peltason Dr, Irvine, CA 92697.

## Abstract

**Background:** Clinic-based estimates of SARS-CoV-2 may considerably underestimate the total number of infections. Access to testing in the US has been heterogeneous and symptoms vary widely in infected persons. Public health surveillance efforts and metrics are therefore hampered by underreporting. We set out to provide a minimally biased estimate of SARS-CoV-2 seroprevalence among adults for a large and diverse county (Orange County, CA, population 3.2 million).

**Methods:** We implemented a surveillance study that minimizes response bias by recruiting adults to answer a survey without knowledge of later being offered a SARS-CoV-2 test. Several methodologies were used to retrieve a population-representative sample. Participants (n=2,979) visited one of 11 drive-thru test sites from July 10^th^ to August 16^th^, 2020 (or received an in-home visit) to provide a finger pin-prick sample. We applied a robust SARS-CoV-2 Antigen Microarray technology, which has superior measurement validity relative to FDA-approved tests.

**Findings:** Participants include a broad age, gender, racial/ethnic, and income representation. Adjusted seroprevalence of SARS-CoV-2 infection was 11.5% (95% CI: 10.5% to 12.4%). Formal bias analyses produced similar results. Prevalence was elevated among Hispanics (vs. other non-Hispanic: prevalence ratio [PR]= 1.47, 95% CI: 1.22 to 1.78) and household income <$50,000 (vs. >$100,000: PR= 1.42, 95% CI: 1.14 to 1.79).

**Interpretation:** Results from a diverse population using a highly specific and sensitive microarray indicate a SARS-CoV-2 seroprevalence of ∼12 percent. This population-based seroprevalence is seven-fold greater than that using official County statistics. In this region, SARS-CoV-2 also disproportionately affects Hispanic and low-income adults.

**Funding:** Orange County Healthcare Agency

## INTRODUCTION

Active and routine surveillance of infectious diseases serves a critical public health function by permitting estimation of overall prevalence and incidence of new infections.^1,2^ California reported the first case of local spread of SARS-CoV-2 causing COVID-19 disease in the US on February 26, 2020 and over the following six months has recorded >586,000 cases and >10,600 deaths from this disease.^3^ Owing in part to limited testing capacity, no state in the US (including California) has enacted routine population-based surveillance of SARS-CoV-2. Instead, as a proxy, states typically collect SARS-CoV-2 polymerase chain reaction (PCR) testing for individual clinical diagnostic purposes. SARS-CoV-2 infection, however, may cause minimal or no symptoms in some individuals.^4,5^ Since persons without (or with minor) symptoms may not seek care, and because testing capacity has frequently been limited, clinic-based estimates may considerably undercount the true incidence of SARS-CoV-2 infections in the population.^6^

Multiple studies have used serologic testing to measure prevalence of COVID-19 in different populations,^4-8^ but the accuracy of these studies has been questioned due to insufficient specificity of the serologic assay for a low prevalence population and due to potential sampling bias.^9,10^ Due to variability in performance of the serologic assay and sampling methodology, these studies have estimated anywhere from a 10-fold to 85-fold higher prevalence as compared to case counts. An ideal seroprevalence study design would recruit a representative sample from the population and detect SARS-CoV-2 (via antibodies from a blood test). To minimize selection bias and skewing of seroprevalence to people with suspected SARS-CoV-2 infection, serum samples would be obtained outside of a clinic *without* oversampling those who are more likely to have been infected (e.g., due to self-selection into a study based on knowledge of symptoms and being offered an antibody test). We know of only one population-based prevalence estimate in the US that meets these criteria.^7^

Orange County (OC) California includes a large, ethnically diverse (34.0% Hispanic, 21.7% Asian) metropolitan region and is the sixth most populous county in the US.^11^ Despite mandating individual and community-wide physical distancing measures, school and business closure, and confinement measures, OC has reported 46,057 cumulative SARS-CoV-2 cases and 957 deaths as of 8/16/20.^12^ We set out to provide a minimally biased estimate of SARS-CoV-2 seroprevalence among all adults in OC.

Our analysis improves upon previous US estimates in several ways.^7,9,13,14^ First, we recruited subjects outside of a clinical setting and employed strategies to minimize bias of recruiting mostly symptomatic cases. Second, we applied a robust SARS-CoV-2 Antigen Microarray technology which has superior sensitivity and specificity relative to what were currently available FDA-approved tests used by others.^15^ Third, we recruited a sufficiently large sample of adults to calculate seroprevalence by race/ethnicity, age, and gender, which may uncover important differences across these groups. Fourth, we recruit subjects by administering a questionnaire *without* initially disclosing an offer for an antibody test. In addition to serving a critical surveillance function, we intend for our results to yield a more accurate measure of the infection fatality risk.

## METHODS

### Recruitment

This study represents a joint effort between University of California, Irvine (UCI) and the Orange County Health Care Agency (OCHCA). We received human subjects approval from the UCI Institutional Review Board (HS# 2020-5952) and obtained informed consent from all study participants.

We focused our study on adults 18 years or older residing in OC on July 1^st^ 2020. We know of no full roster of the adult population in OC that would permit sampling based on complete enumeration. Absent this information, we used a proprietary database, reflecting the age, income, and racial/ethnic diversity of OC and maintained by SoapBoxSample, an LRW Group Company, to recruit participants. This proprietary database contains contact information for 800,000 adults—almost one-third of all adults in OC. The database, moreover, contains sociodemographic information which allows us to assess potential non-response bias.

Using this database, we invited (via email [36.4%] or random-digit telephone dialing [63.6%]) one resident per household to participate in a study about their opinions of COVID-19, without initial mention of SARS-CoV-2 antibody testing. We did not allow the opportunity to defer the survey to another household member. Participants completed a survey regarding socio-demographics (e.g., age, gender, race/ethnicity, household income), daily work and social activities related to SARS-CoV-2, any known previous infection with SARS-CoV-2, and history of SARS-CoV-2 symptoms in the last few months. Once the respondent provided these answers, we then asked if they were willing to participate in a drive-thru finger-prick blood test for SARS-CoV-2 antibodies. Participants received a $10 gift card as compensation for completion of the survey and blood test.

We aimed to reach specific quotas for enrollment for demographic subgroups. Based on US Census-derived population distributions in OC of age, gender, race/ethnicity, and household income,^11^ we targeted recruitment to ensure adequate sample size of subjects from the following strata: age-by-gender (18 to 34 years, 35 to 54 years, 55 years or above; by male, female), race/ethnicity (Hispanic, Asian non-Hispanic, and other non-Hispanic including white and Black, and household income (<$50,000, $50,000-99,999, and $100,000 or above).

We encouraged participation of racial/ethnic minorities and/or lower-income adults in several ways. First, for four of the six weeks of the survey, we offered the survey in the five most commonly spoken languages in OC (i.e., English, Spanish, Mandarin, Vietnamese, and Korean). Second, we invited all household members of underrepresented groups to receive testing at the drive-thru site. Third, for the last two weeks of the survey we targeted particular zip codes with a large proportion of racial/ethnic minorities. To increase participation among persons considered vulnerable (e.g., aged >65 years), we offered in-home visits at which licensed phlebotomists obtained the finger-prick blood sample at the participant’s home.

For participants who agreed to the antibody test, we invited them to one of eleven drive-thru test sites that spanned the geography of OC. These sites were distributed across OC to permit ease of access (by public transit and/or less than 15 minutes driving time) and previously identified by OC as Points of Dispensing sites as allowing ease of drive-thru testing given existing security and experience with large numbers of people. We assigned each participant a unique alphanumeric code, to be presented at the drive-thru test site, to ensure that only survey participants (and not others interested in receiving the antibody test) were processed. To increase the participation rate, we sent appointment reminders via email and telephone at regular intervals leading up to the scheduled date and time. We conducted SARS-CoV-2 antibody testing from July 10^th^ to August 16^th^, 2020 on Thursdays through Sundays in an attempt to accommodate a range of participant work schedules and availability. We used UCI student and alumni volunteers to staff the sites overseen by trained faculty.

### Laboratory Methods

We assessed past SARS-CoV-2 infection using a coronavirus antigen microarray (CoVAM). This microarray approach has been widely used for multiple pathogens.^16^ Unlike then available FDA authorized tests which detect antibodies against one or two antigens,^17^ the CoVAM quantitatively measures IgG and IgM antibodies against 12 antigens from SARS-CoV-2, including spike (S) as whole protein or separated domains including S1, S2, and receptor-binding domain (RBD), nucleocapsid protein (NP), and membrane (M) protein (**Supplemental Material eFigure 1**). The CoVAM also measures IgG and IgM antibodies against 53 antigens from other viruses that cause acute respiratory illness, including SARS-1, MERS, and the four seasonal coronaviruses, which can be used for assessment of antibody cross-reactivity. The antigens are printed onto microarrays in quadruplicate, probed with blood specimens and secondary antibodies for IgM and IgG, and imaged to determine background-subtracted median fluorescence intensity averaged across quadruplicate spots as previously described.^15,16,18,19^ The protein microarray requires less than 100µl of blood, which we retrieved from participants using a finger-prick for blood collection into capillary tubes. We processed blood specimens for plasma separation and microarray analysis in a high throughput manner within four days of collection.

### Statistical Analysis

SARS-CoV-2 antibody reactivity was determined as previously described.^15^ Briefly, the CoVAM data for each specimen was compared to 99 positive PCR-confirmed cases with blood collected at least 7 days (range 7-50 days, median 11 days) post symptom onset and 88 negative pre-pandemic controls with blood collected prior to November 1, 2019. From the 12 SARS-CoV-2 antigens on the array, a linear regression model was trained on positive and negative controls to determine optimal weighted combinations of reactive antigens to calculate composite SARS-CoV-2 IgG and IgM antibody levels that discriminate these two groups. Based on Receiver Operating Characteristic curve analyses, reactivity thresholds for IgG and IgM were chosen to classify unknown specimens demonstrating 100% specificity and 94% sensitivity for either IgM or IgG.^15,18^ These test characteristics compare favorably to current FDA-authorized test kits^17^ and indicate a very low likelihood of false positive detection of SARS-CoV infection.

We first calculated crude seroprevalence for SARS-CoV-2 antibodies (excluding household members of participants who were tested after study recruitment) by dividing the number of reactive cases by the total number of participants. Next, we estimated population-representative SARS-CoV-2 seroprevalence by post-stratifying results by the age, gender, race/ethnicity, and income distribution for each of the 88 zip codes in OC. This process, known as raking,^20^ uses age-gender-race/ethnicity population estimates and income population estimates from the US Census^11^ to determine appropriate weights for estimating the overall seroprevalence for OC as a whole. Participants who did not provide complete demographic information were excluded from the weighted analyses, as were empty strata. In secondary analyses, we used multiple imputation to assess non-response bias and Bayesian methods to adjust for antibody test accuracy (**eTable 2, Supplemental Material**).

We estimated prevalence ratios (i.e., relative risk across subgroups of a seropositive SARS-CoV-2 test) and 95% uncertainty intervals using both unweighted and population-weighted^21^ log-binomial multiple regression, comparisons across age, gender, income, and race/ethnicity subgroups, mutually adjusting for these covariates as well as sampling week, to limit confounding by any changes in recruitment and seroprevalence over time. We determined reference groups based on the stratum with the largest sample size. Finally, we estimated the infection fatality risk among adults in OC by dividing the number of cumulative excess SARS-CoV-2 deaths among adults by the number of implied past infections (i.e., seroprevalence × adult population) at the time of the survey that could have led to a recorded death by the time of our analysis. We compared this infection fatality risk to that using published statistics from OCHCA.^12^

## RESULTS

Over the study period, 4,555 adults completed the survey and consented to participate in the blood test. Of these participants, 2,979 (65.4%) showed up to provide a viable blood sample for CoVAM analysis from July 10^th^ to August 16^th^, 2020. **Figures 1A and 1B** illustrate negative and positive SARS-CoV-2 test results using CoVAM. Comparison of the black circles (i.e., patient IgG and IgM values) across these figures show clear differences in the SARS-CoV-2 antibody reactive profile between a negative and positive test. A full heat map of all specimens, with classification and positive and negative controls, appears in **Supplemental Material (eFigure 1**).

**Figures 1A and 1B.**
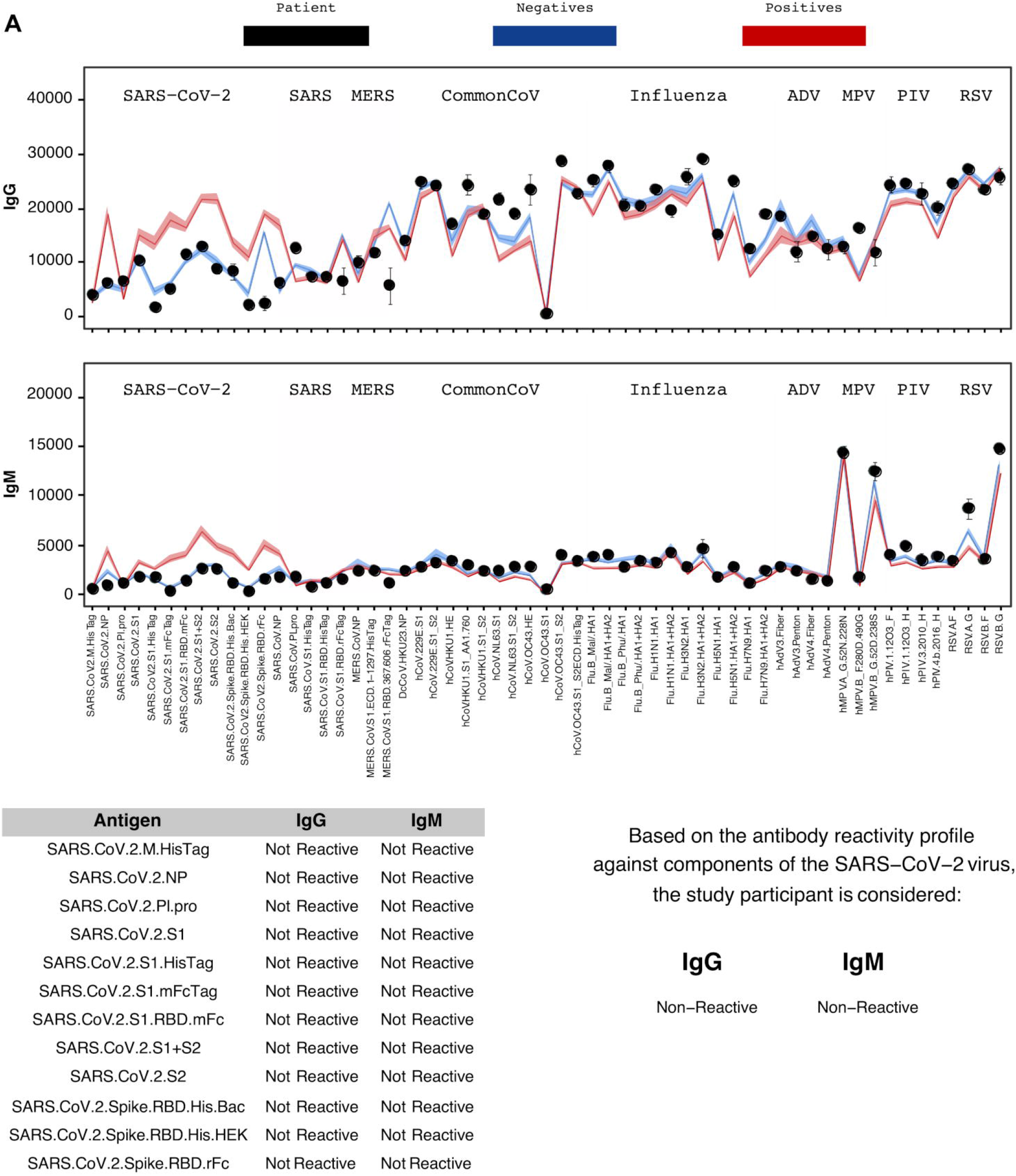
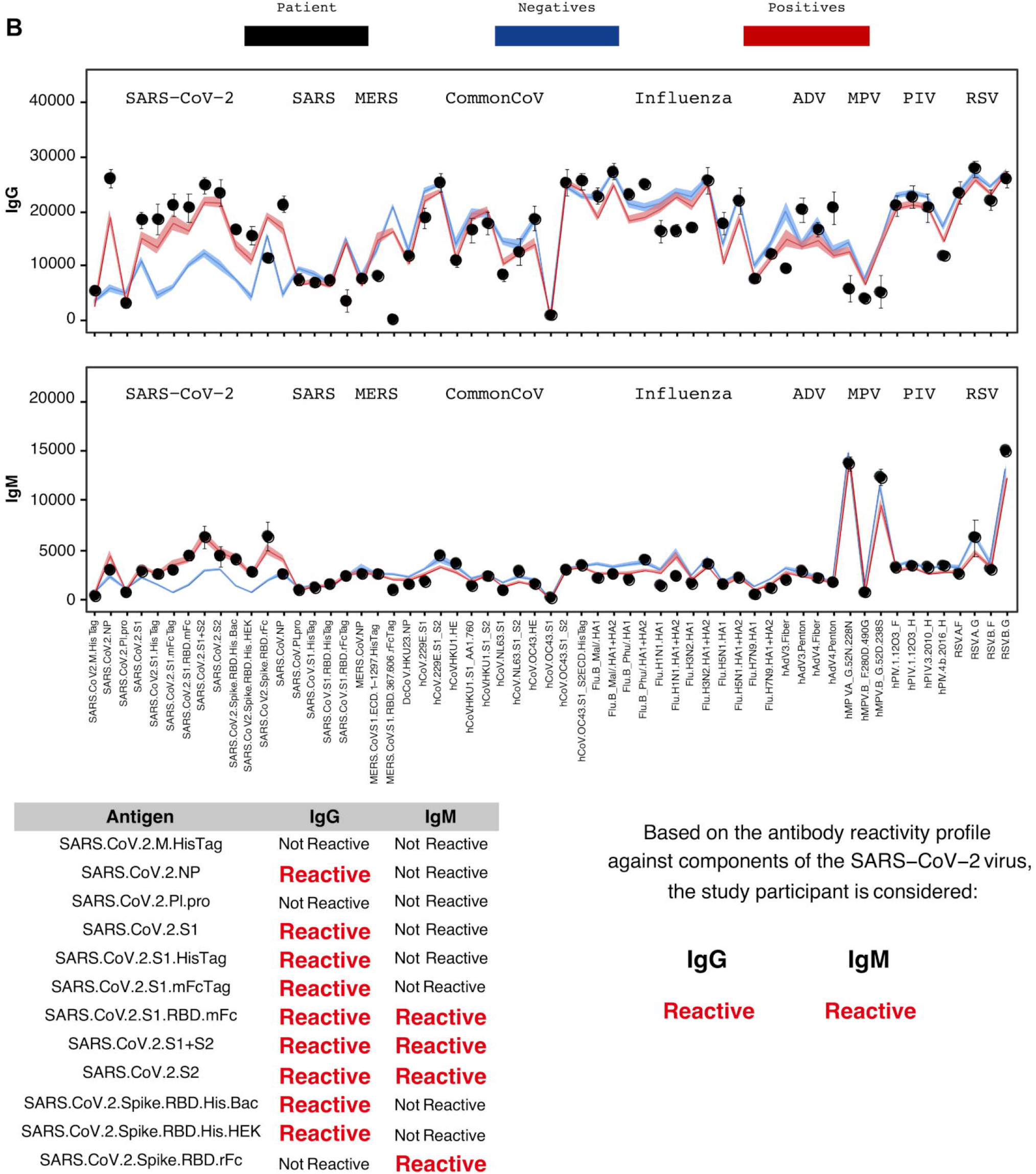
Coronavirus antigen microarray result report for representative negative (A) and positive (B) study participants. For each result, the IgG and IgM antibody reactivity quantified by mean fluorescence intensity is plotted with comparison to reference positives and negatives and classified as “Reactive” or “Non-Reactive” for each individual antigen and overall for IgG and IgM.

Our recruitment strategy successfully garnered participation from a broad range of socioeconomic, age, and racial/ethnic backgrounds (**Table 1**). The overall unadjusted seroprevalence of SARS-CoV-2 is 11.8% (351/ 2,979). **Table 1** also shows elevated crude SARS-CoV-2 seroprevalence among Hispanic and low-income (<$50,000) adults.

**Table 1:** **Unadjusted** SARS-CoV-2 seroprevalence by social, demographic and survey characteristics among 2,979 adults in Orange County, CA, July 10^th^ to August 16^th^, 2020.

| Characteristic | Participants |  | Seroprevalence |  |
| --- | --- | --- | --- | --- |
|  | N | (%) | Cases/total | (%) |
| Overall | 2979 | (100) | 351/2979 | (11.8) |
| **Age** |  |  |  |  |
| 18 to 34 years | 673 | (22.6) | 83/673 | (12.3) |
| 35 to 54 years | 1362 | (45.7) | 168/1362 | (12.3) |
| ≥55 years | 944 | (31.7) | 100/944 | (10.6) |
| **Gender** |  |  |  |  |
| Female | 1680 | (56.4) | 223/1680 | (13.3) |
| Male | 1296 | (44.6) | 128/1296 | (9.9) |
| **Race/Ethnicity** |  |  |  |  |
| Hispanic (including multi-race) | 1108 | (37.2) | 172/1108 | (15.5) |
| Asian (Non-Hispanic, Non-African American) | 469 | (15.7) | 50/469 | (10.7) |
| Other Non-Hispanic (including white and African American) | 1402 | (47.1) | 129/1402 | (9.2) |
| **Household Income per year** |  |  |  |  |
| <\$50,000 | 655 | (22.0) | 98/655 | (15.0) |
| \$50,000 to \$99,999 | 722 | (24.2) | 96/722 | (13.3) |
| ≥\$100,000 | 1090 | (36.6) | 87/1090 | (8.0) |
| Prefer not to answer | 512 | (17.2) | 70/512 | (13.7) |
| **Had contact with anyone suspected or confirmed of COVID-19?** |  |  |  |  |
| Yes | 579 | (19.4) | 120/579 | (20.7) |
| No | 1850 | (62.1) | 191/1850 | (10.3) |
| Don't know | 546 | (18.3) | 40/546 | (7.3) |
| **Had any SARS-CoV-2 symptoms in last 2 months?** |  |  |  |  |
| Yes | 1038 | (34.8) | 166/1038 | (16.0) |
| No | 1941 | (65.2) | 185/1941 | (9.5) |

The fact that our sample skews towards higher-income earners and older adults (**Supplemental Material eTable 3**) indicates that post-stratification weighting may provide a more representative estimate of SARS-CoV-2 seroprevalence for OC. This weighting produced an adjusted seroprevalence of 11.5% (95% Confidence Interval: 10.5% to 12.4%). We also performed sensitivity analyses to ascertain potential bias induced by differential non-response of adults with respect to SARS-CoV-2 positivity status. We assessed non-response bias in several ways (**Supplemental Material, Bias Analysis**). Depending on the level of differential non-response by persons suspected to be SARS-CoV-2 positive and the adjustment strategy used, adjusted point estimates for seroprevalence range from 11.3% to 14.4%.

Consistent with the unadjusted results, adjustment for post-stratification weighting indicates that prevalence is elevated among Hispanics (vs. other non-Hispanic: PR= 1.47, 95% CI: 1.22 to 1.78), household income <$50,000 (vs. >$100,000: PR= 1.42, 95% CI: 1.14 to 1.79), and symptomatic adults (vs. no symptoms: PR= 1.68, 95% CI: 1.41 to 2.00; **Table 2**). Males also show a lower prevalence (PR= 0.85 95% CI: 0.71 to 1.01) relative to females.

**Table 2:**
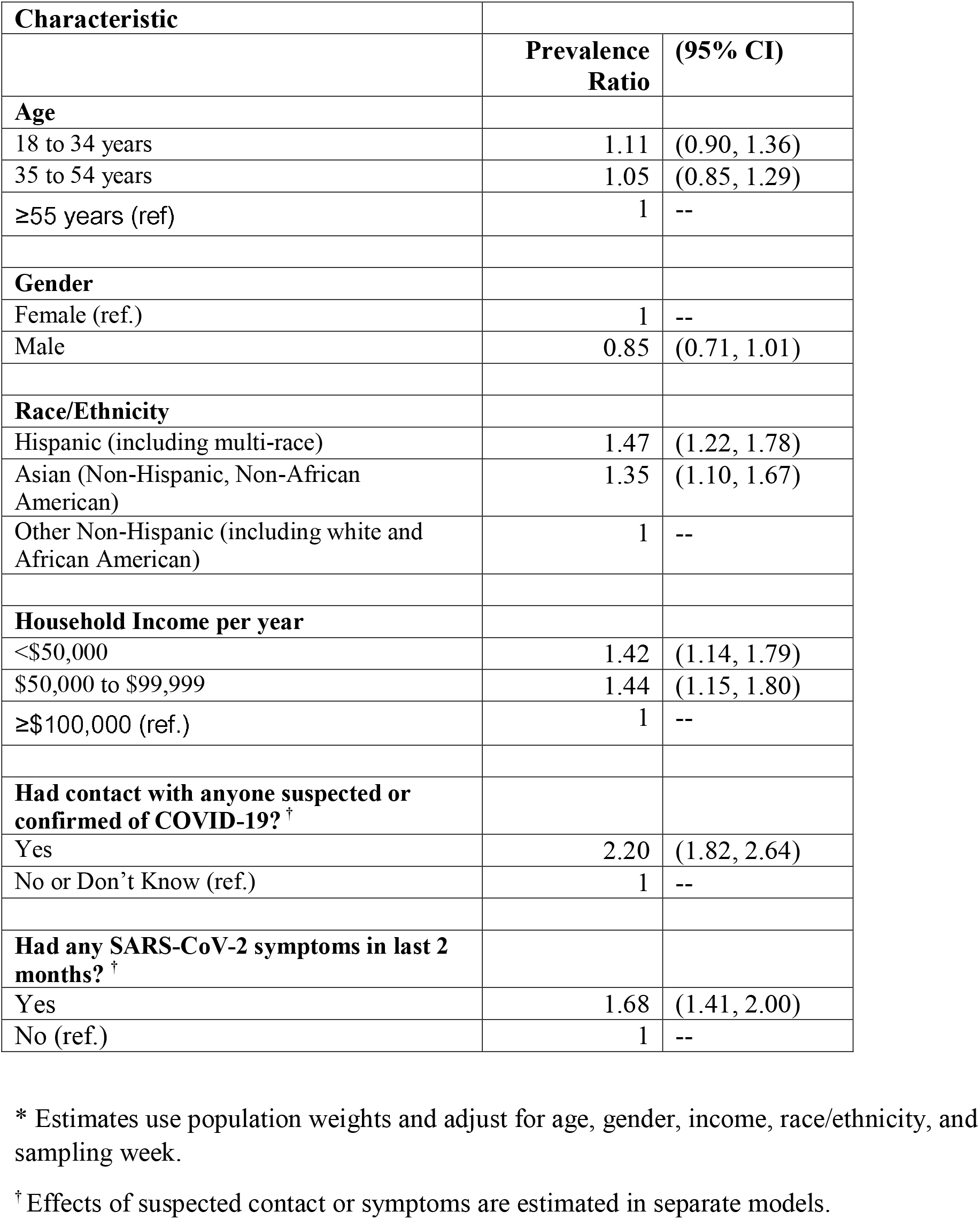
**Adjusted\*** SARS-CoV-2 seroprevalence ratios among adults in Orange County, CA, July 10^th^ to August 16^th^, 2020,

| **Characteristic** | **Prevalence Ratio** | **(95% CI)** |
| --- | --- | --- |
| **Age** |  |  |
| 18 to 34 years | 1.11 | (0.90, 1.36) |
| 35 to 54 years | 1.05 | (0.85, 1.29) |
| ≥55 years (ref) | 1 | -- |
| **Gender** |  |  |
| Female (ref.) | 1 | -- |
| Male | 0.85 | (0.71, 1.01) |
| **Race/Ethnicity** |  |  |
| Hispanic (including multi-race) | 1.47 | (1.22, 1.78) |
| Asian (Non-Hispanic, Non-African American) | 1.35 | (1.10, 1.67) |
| Other Non-Hispanic (including white and African American) | 1 | -- |
| **Household Income per year** |  |  |
| <\$50,000 | 1.42 | (1.14, 1.79) |
| \$50,000 to \$99,999 | 1.44 | (1.15, 1.80) |
| ≥\$100,000 (ref.) | 1 | -- |
| **Had contact with anyone suspected or confirmed of COVID-19? <sup>†</sup>** |  |  |
| Yes | 2.20 | (1.82, 2.64) |
| No or Don't Know (ref.) | 1 | -- |
| **Had any SARS-CoV-2 symptoms in last 2 months? <sup>†</sup>** |  |  |
| Yes | 1.68 | (1.41, 2.00) |
| No (ref.) | 1 | -- |

\* Estimates use population weights and adjust for age, gender, income, race/ethnicity, and sampling week.
<sup>†</sup> Effects of suspected contact or symptoms are estimated in separate models.

We calculated the incidence fatality risk, separately for adults 18 to 64 years and those 65 years and above, given the strong age-related morbidity and mortality of SARS-CoV-2 (**Supplemental Material, Incidence Fatality Risk**).^22^ Using the seroprevalence results gives a range of infection fatality rates that are 0.94 to 1.9 deaths per 1,000 persons for adults 18 to 64 years, and 10.5 to 11.6 deaths per 1,000 persons for adults 65 years and above. In comparing these values to those calculated using only reported statistics (through to 8/16/20, the end of our study period) from OCHCA, for 18 to 64 year olds the population-based SARS-CoV-2 infection fatality rate is an estimated 3.2 to 6.3 fold lower. Also, for 65 years and above, the population-based SARS-CoV-2 infection fatality rate is an estimated 9.0 to 10.0 fold lower than that reported by OCHCA.

## DISCUSSION

We set out to provide a minimally biased estimate of SARS-CoV-2 seroprevalence among adults in Orange County, a densely-populated and diverse county in southern California. We recruited subjects outside of a clinical setting and without their knowledge that we would ultimately offer an antibody test. Results from a socioeconomically and demographically diverse population of >2,900 adults using a highly specific and sensitive CoVAM microarray indicate a SARS-CoV-2 seroprevalence of approximately 12 percent. This SARS-CoV-2 point seroprevalence among adults for July to August 2020 is seven-fold greater than the cumulative incidence of diagnosed cases reported to Orange County.

Our seroprevalence estimate is greater than that reported from antibody surveys among blood donors nationwide,^23^ but less than that reported from New York City.^14,24^ Early community serosurveys in California ranged from 4.65% among adults in neighboring LA^7^ to 2.8% among adults and children in Santa Clara County,^25^ both of which suggested infection was substantially more widespread in those regions than case counts implied. Other states have similarly estimated prevalence between 2 and 4%.^9,26^ We, however, caution against direct comparisons given the different course, sample (clinic vs. community) and timing of the epidemic across regions, and different sensitivity and specificity of test used.

The seroprevalence of SARS-CoV-2 in OC is 1.8 fold greater among Hispanics than among the referent group of mostly non-Hispanic whites. Previous clinic-based research in Baltimore/Washington D.C.^27^ and Orange County^28^ also supports this finding. Greater prevalence of SARS-CoV-2 among Hispanics may arise from work in settings that do not allow for physical distancing, work under hazardous conditions due to economic necessity, and residence in relatively dense housing conditions.^28-31^

Strengths of our study include the recruitment of a large sample of adults without their knowledge that we would ultimately offer an antibody test, which minimizes response bias. We also recruited a diverse population in multiple languages which permits precise estimation of SARS-CoV-2 seroprevalence by age, gender, race/ethnicity, and income subgroups. The detailed survey, moreover, allowed us to assess the sensitivity of main results to biased participation in blood testing among symptomatic individuals. The CoVAM microarray technology is a study strength in that it quantifies antibody responses to a wide range of SARS-CoV-2 proteins. The specificity and sensitivity of CoVAM compares favorably to other antibody test kits used to estimate seroprevalence of past infection in the US. In addition, the lack of reliance of CoVAM on any one specific SARS-CoV-2 protein (e.g., the “N” protein) minimizes the risk of a false negative test resulting from inter-individual variability in antibody profiles across antigens. CoVAM also requires only a few microliters of blood, which is practical for increasing compliance during large surveillance programs.

We did not randomly sample the entire adult population in OC owing to the logistical constraint of having neither a full roster of residents nor complete demographic information on individuals in the commercially obtained database. In addition, over a third of respondents who consented did not provide a blood test. Although this circumstance *<u>per se</u>* does not introduce bias,^32-34^ we cannot rule out the possibility of differential nonresponse with respect to past SARS-CoV-2 infection. Our analyses, however, permit evaluation of the potential role of bias, suggests minimal bias in estimates, and makes explicit our assumptions, thereby permitting replication (**Supplemental Material, Bias Analysis**).

Whereas we retrieved blood samples over a span of six weeks, relatively few samples for any particular week precluded calculations over time of SARS-CoV-2 seroprevalence precisely when OC reported an increase in reported cases. In addition, CoVAM detects past infection by quantifying antibodies that respond to a broad set of SARS-CoV-2 proteins, but it may fail to detect a small subset of new infections that have not yet elicited a sufficient antibody response. We also did not examine children <18 years of age. Finally, findings pertain to OC only—an area thought to be affected earlier by SARS-CoV-2 than many other US regions, but not as affected as New York City—and should not be used to generalize to other places or times.

The implications of our study are three-fold. First, the widespread seroprevalence of SARS-CoV-2 in OC warrants continued public health measures of physical distancing, proper and consistent use of face masks, ventilation, and hand hygiene. As has been reported elsewhere, SARS-CoV-2 infections are underreported; our seroprevalence survey suggests that infections are underreported by a factor of 7.6. Second, in addition to contact tracing, authorities may want to consider active surveillance of novel infections. This active surveillance would involve both a population-based component (e.g., 800-1,000 tests per week in a representative sample of county or city residents) as well as a targeted component on higher-risk groups or places (e.g., specific census tracts, nursing home residents, or laborers in high-density settings). Such surveillance, unlike clinic- or hospital-based strategies, would provide a less-biased estimate of the rate of new SARS-CoV-2 infections. Third, our estimates of infection fatality rates for adults 18 to 64 years and those 65 years and above is several-fold lower than that using county reports but lies within the range of other fatality risk studies from other US regions.^35^ This updated estimate should inform the broader policy debate in the US regarding the relative benefits and limitations of various SARS-CoV-2 mitigation strategies.

## Supporting information

Supplemental_Material_CoVAM_map

## Data Availability

These data contain personal identifying health information and are therefore not available for public use or dissemination. However, researchers interested in accessing a de-identified version of the dataset may contact the corresponding Author. In addition, the R-program code for the data analysis are available in the Supplemental Material.

## Acknowledgments

We are grateful to Dr. Neeraj Sood at the University of Southern California Price School of Public Policy for advice regarding study design, and for providing us the survey instrument used in their Los Angeles, CA seroprevalence study in conjunction with Dr. Paul Simon at the Los Angeles County Department of Public Health.

**Supplemental Material, eFigure 1**. Heat map of IgM (A) and IgG (B) antibodies by coronavirus antigen microarray. Each column represents antibody responses in a single individual, arranged from left to right as reference positives, study positives and negatives as determined by generalized linear model (glm), and reference negatives with color-coded labels above. Each row represents antibody responses against a single antigen, arranged from top to bottom as SARS-CoV-2 antigens, other coronavirus antigens, and other respiratory virus antigens with color-coded labels to the left (ADV = adenovirus, MPV = metapneumovirus, PIV = parainfluenza virus, RSV = respiratory syncytial virus). The antibody reactivity against each antigen is quantified by mean fluorescence intensity (white = low, black = medium, red = high).

