## Supplemental_Material_Text for "Estimated Seroprevalence of SARS-CoV-2 Antibodies Among Adults in Orange County, California"

**Bias Analysis**

Although survey researchers disagree regarding the best way to analyze survey data collected using quota sampling, common choices include unweighted analysis based on simple random sampling or weighted post-stratification, with the understanding that confidence intervals may not be accurate and should be interpreted with caution.^1,2^ We employ both methods here, as well as multiple imputation to adjust for non-response in antibody testing after survey completion, and Bayesian adjustment for antibody test accuracy.

For weighted analysis, multiply-stratified population estimates were obtained from the 2018 American Community Survey and other US Census surveys^3^ for each combination of gender (male or female), race/ethnicity (Hispanic, Asian, or white/other non-Hispanic), age (18-34, 35-54, or 55+), and zip code (87 unique zip codes) in OC, and for each combination of annual income (<$50,000, $50,000-$99,000, or $100,000+) and zip code. Population counts were not available multiply-stratified on all variables targeted in the quota sample, so we used raking (iterative post-stratification by each set of population counts) to combine the overlapping population counts.^4^ For the weighted analyses, participants were excluded if they were recruited as a household member, if they did not provide all of the post-stratification variables, or if stratified population counts were not available for their demographic category (e.g., non-binary gender, or preferred not to answer question on household income). In total, sufficient information was available to include 2,454 participants with valid antibody test results (280 of whom tested positive) in the weighted analyses. All weighted analyses were conducted using the *survey* library in R. Code is available at the end of this supplement.

Some participants completed most or all of the questionnaire, but did consent to the blood draw and/or did not provide a valid blood sample. In total, 9,313 non-household participants completed the symptom questionnaire and provided a valid zip code. Unweighted characteristics for these 9,313 participants and the 2,454 with valid antibody test results are compared in **eTable 1**. Those who provided complete demographic information and a blood sample were substantially more likely to have COVID-19 symptoms, to be male, to be Hispanic or Asian, or to have an annual income <$50,000 than those who did not provide a blood sample. In order to assess the potential impacts of non-response bias with regard to providing a blood sample, we performed multiple imputation of missing values of COVID-19 status, income, and other key variables. We used a limited set of variables for the imputation model, in order to avoid singularities and other computational difficulties: antibody status, race/ethnicity, age, gender, income, week of blood sample collection, zip code, known or suspected contact with a COVID-19 positive individual, self-reported symptoms, primary language, current employment status, working at home or outside of the home, and perceived likelihood of having been infected with COVID-19. We used 25 imputations from a fixed seed, generated by the *mice* library in R (version 3.6.3). For weighted analyses with multiple imputation, the same raking method and survey weighting procedure was used for each imputed data set. Rubin's rules were used to combine estimates across imputed datasets.^5^

To account for imperfect sensitivity and specificity of the antibody test, we performed Bayesian analyses accounting for the known relationship^6^ between the probability of a randomly sampled person testing positive, Pr(T+), and the prevalence of past COVID-19 infection, Pr(D+):

Pr(T+) = Pr(T+|D+) * Pr(D+) + Pr(T+|D-) * Pr(D-)

= sensitivity * prevalence + (1 - specificity) * (1 - prevalence)

We used a binomial likelihood function for observed test positivity for unweighted analyses, and a normal likelihood function for seroprevalence estimates from weighted analyses. The results from our 99 positive controls and 88 negative controls were used to establish beta-binomial priors for the antibody test sensitivity and specificity, with parameters *k*+1 and *n*-*k*+1 where for each group of controls *n* is the number tested and *k* is the number correctly classified, and we used a diffuse unif(0,1) prior for the prevalence. Other diffuse priors for the prevalence produced similar results. Each Bayesian analysis was performed using JAGS and the *rjags* library in R, using 3 independent Markov chains with well dispersed initial values and 10,000 iterations each, after 5,000 iterations in adaptive mode. The multivariate and univariate potential scale reduction factors and their upper confidence limits were 1.02 or less for each analysis, indicating good convergence.^7^

Results for the various adjustment methods are shown in **eTable 2**.

**eTable 1. Characteristics of participants in weighted analyses and multiple imputation**

| **Variable** | **Level** | **Weighted**  **(n = 2454)** | **Multiple Imputation**  **(n = 9313)** |
| --- | --- | --- | --- |
| Age | 18-34 years | 574 (23%) | 2349 (25%) |
|  | 35-54 years | 1100 (45%) | 3694 (40%) |
|  | 55+ years | 780 (32%) | 3270 (35%) |
| Race/ethnicity | Hispanic | 1210 (49%) | 3999 (43%) |
|  | Asian | 840 (34%) | 1376 (15%) |
|  | white/other non-Hispanic | 404 (16%) | 3938 (42%) |
| Gender | Male | 1372 (56%) | 3980 (43%) |
|  | Female | 1082 (44%) | 5314 (57%) |
|  | Nonbinary/Missing | - | 19 (<1%) |
| Income | <$50,000 | 1084 (44%) | 2833 (30%) |
|  | $50,000-$99,000 | 649 (26%) | 2008 (22%) |
|  | $100,000+ | 721 (29%) | 2551 (27%) |
|  | Missing / Prefer not to state | - | 1921 (21%) |
| COVID-19 symptoms | Yes | 1559 (64%) | 2641 (28%) |
|  | No | 895 (36%) | 6672 (72%) |

**eTable 2. Seroprevalence estimates for weighted and alternative analyses**

| **Method** | **Point estimate and 95% CI*** |
| --- | --- |
| Weighted, only those with valid test results | 11.5% (10.5%, 12.4%) |
| Unweighted, with multiple imputation | 14.4% (12.9%, 15.9%) |
| Weighted, with multiple imputation | 13.3% (11.9%, 14.6%) |
| Unweighted, with Bayesian adjustment | 11.6% (8.6%, 13.7%) |
| Weighted, with Bayesian adjustment | 11.3% (8.1%, 13.2%) |
| Weighted, with multiple imputation and Bayesian adjustment | 13.3% (9.9%, 15.5%) |

* For each frequentist analysis, the point estimate and 95% Confidence Interval are provided. For each Bayesian analysis, the posterior mean and 95% Credible Interval are provided.

**R code for Weighted Analysis	Author: Scott Bartell	Date: 9/20/20**

library(survey)

library(mice)

library(mitools)

library(miceadds)

library(rjags)

load("cov18noid.rdata")

\# popwt1, popwt3, popwt3 are estimated population sizes, by zipcode, of age/sex/race categories

\# wiu50k, wi5099k, wio100k are estimates population count by zip code, in each income category

\# DV_IMPORT_TEST_RESULTresult (blood test result, reactive, non reactive, fail, not recorded)

\# E4 is 1 if blood sample provided

\# exclude household recruits (DV_PANEL is 7)

\# exclude anyone not completing Q13r9 (symptom survey)

\# Q1 is gender (1 for male, 2 for female, 3 for nonbinary)

\# Q2 is age (cats 2-4 for age 18-34, 5-8 for 35-54, 9-15 for 55+)

\# Q2HIDDEN_AGEBUCKET already collapsed (1 for age 18-34, 2 for 35-54, 3 for 55+)

\# Q6r1 through Q6r6 are race/ethnicity

\# Q6HIDDEN_QUOTA is race/ethnicity group (1 for Hispanic, 2 for Asian, 3 for other)

\# Q7HIDDEN_QUOTA is income group (1 for <50k, 2 for inc50t99k, 3 for 100K+, 4 for NA)

\# check to make sure everyone with a valid test result completed the symptom survey

any(is.na(cov18noid$Q13r9) & cov18noid$DV_IMPORT_TEST_RESULTresult %in% c("R","N") )

\# exclusions: household recruits (DV_PANEL 7) and those who didn't provide symptoms (Q13r9)

d1 = cov18noid[cov18noid$DV_PANEL != 7 & !is.na(cov18noid$Q13r9),]

\# clean and convert key variables to factors

d1$covid = factor(d1$DV_IMPORT_TEST_RESULTresult)

levels(d1$covid) = c(NA,NA,"N",NA,"R")

d1$covid19 = as.numeric(d1$covid=="R")

d1$age = factor(d1$Q2HIDDEN_AGEBUCKET)

levels(d1$age) = c("18-34","35-54","55+")

d1$race = factor(d1$Q6HIDDEN_QUOTA)

levels(d1$race) = c("hispanic","asian","other")

d1$gender = factor(d1$Q1)

levels(d1$gender) = c("male","female",NA)

d1$zip = factor(d1$wiu50k)

d1$income = factor(d1$Q7HIDDEN_QUOTA)

levels(d1$income) = c("<50K","50-99K","100K+",NA)

d1$dateweek = factor(d1$dateweek)

d1$contact = factor(d1$Q11)

levels(d1$contact) = c("Yes","No or Don't Know","No or Don't Know",NA)

d1$symptoms = factor(d1$Q13HIDDEN_2)

levels(d1$symptoms) = c("Yes","No")

d1$language = factor(d1$DV_LANG)

d1$employed = factor(d1$Q6d)

d1$workathome = factor(d1$Q6f)

d1$sector = factor(d1$Q6g)

d1$likely = factor(d1$Q19)

\# make a new variable for income-zip population estimates

d1$popwt4 =	(d1$income=="<50K") * d1$wiu50k +

(d1$income=="50-99K") * d1$wi5099k +

(d1$income=="100K+") * d1$wio100k

\#### Unweighted model

mean(d1$covid == "R", na.rm=T)

\####	Weighted Model

\# post-stratification only, raking on income-zip and gender-age-race-zip

\# excluding NAs

\# exclude those with no antibody test results

\# construct marginal tables using popwt3 and popwt4

dw = d1[!is.na(d1$age) & !is.na(d1$race) & !is.na(d1$gender) & !is.na(d1$income) &

!is.na(d1$zip) & !is.na(d1$popwt3) & !is.na(d1$popwt4) &!is.na(d1$covid), ]

dw$race = factor(dw$race,levels(dw$race)[c(3,1:2)])

dw$gender = factor(dw$gender,levels(dw$gender)[2:1])

dw$age = factor(dw$age,levels(dw$age)[c(3,1:2)])

dw$income = factor(dw$income,levels(dw$income)[c(3,1:2)])

dw$contact = factor(dw$contact,levels(dw$contact)[2:1])

dw$symptoms = factor(dw$symptoms,levels(dw$symptoms)[2:1])

dw$strata1 = factor(paste(dw$age,dw$race,dw$gender,dw$zip,sep="-"))

dw$strata2 = factor(paste(dw$income,dw$zip,sep="-"))

strata1marg = dw[!duplicated(dw[,"strata1"]),c("strata1","popwt3")]

strata2marg = dw[!duplicated(dw[,"strata2"]),c("strata2","popwt4")]

names(strata1marg)[2] = "Freq"

names(strata2marg)[2] = "Freq"

\# SRS design, then post-stratify independently and on both marginals using raking

design1 = svydesign(id=~1, data=dw)

design1rake = rake(design1, sample=list(~strata1,~strata2),

population=list(strata1marg,strata2marg), control=list(maxit=50, epsilon=.01))

\# estimate seroprevalence

(m0 = svymean(~covid19, design1rake, deff=T))

confint(m0)

t.test(dw$covid == "R") # compare results to unweighted analysis (or use binom.test)

\# Entries for Table S1

summary(dw$age)

summary(dw$age) / sum(summary(dw$age))

summary(dw$race)

summary(dw$race) / sum(summary(dw$race))

summary(dw$gender)

summary(dw$gender) / sum(summary(dw$gender))

summary(dw$income)

summary(dw$income) / sum(summary(dw$income))

summary(dw$symptoms)

summary(dw$symptoms) / sum(summary(dw$symptoms))

\# stratified by design variables

(p.race = svyby(~covid19, ~race, svymean, design=design1rake,deff=T))

(p.gender = svyby(~covid19, ~gender, svymean, design=design1rake,deff=T))

(p.age = svyby(~covid19, ~age, svymean, design=design1rake,deff=T))

(p.income = svyby(~covid19, ~income, svymean, design=design1rake,deff=T))

\# svyby(~covid, ~zip, svymean, design=design1rake,deff=T)

\# stratified by week

svyby(~covid19, ~dateweek, svymean, design=design1rake,deff=T)

rbw = svyby(~covid19, ~race+dateweek, svymean, design=design1rake)

barplot(rbw)

\# multiple log binomial regression for RRs

\# default log-binomial models not converging; use quasipoisson approximation for starting values

\# key confounders

m1start = svyglm(covid19~race+age+gender+income+dateweek,

design=design1rake, family=quasipoisson(log)) # get starting values from qp model

m1 = update(m1start,family=quasibinomial(log),start=m1start$coef)

round(exp(cbind(est=m1$coef,confint(m1))),2)	# pt est and CI, weighted

\# including intermediates

m2start = svyglm(covid19~contact + race+age+gender+income+dateweek,

design=design1rake, family=quasipoisson(log))

m2 = update(m2start,family=quasibinomial(log),start=m2start$coef)

round(exp(cbind(est=m2$coef,confint(m2))),2)	# pt est and CI, weighted

m3start = svyglm(covid19~symptoms + race+age+gender+income+dateweek,

design=design1rake, family=quasipoisson(log))

m3 = update(m3start,family=quasibinomial(log),start=m3start$coef)

round(exp(cbind(est=m3$coef,confint(m3))),2)	# pt est and CI, weighted

\# with MI

\# imp <- mice(d1)	# singularities; reduced variable space to key variables

\# zip code has too many categories (>50) to impute missing values

\# including work sector generates too many weights

d1key <- d1[!is.na(d1$zip),

c("covid","race","age","gender","income","dateweek","zip","contact","symptoms",

"language","employed","workathome","likely")]

dim(d1key)	# get sample size

\# Entries for Table S1

summary(d1key$age)

summary(d1key$age) / sum(summary(d1key$age))

summary(d1key$race)

summary(d1key$race) / sum(summary(d1key$race))

summary(d1key$gender)

summary(d1key$gender) / sum(summary(d1key$gender))

summary(d1key$income)

summary(d1key$income) / sum(summary(d1key$income))

summary(d1key$symptoms)

summary(d1key$symptoms) / sum(summary(d1key$symptoms))

\# obtain population totals by stratum using entire dataset

d1marg <- within(d1, {

strata1 = factor(paste(age,race,gender,zip,sep="-"))

strata2 = factor(paste(income,zip,sep="-"))

})

strata1marg = na.omit(d1marg[!duplicated(d1marg[,"strata1"]),c("strata1","popwt3")])

strata2marg = na.omit(d1marg[!duplicated(d1marg[,"strata2"]),c("strata2","popwt4")])

names(strata1marg)[2] = "Freq"

names(strata2marg)[2] = "Freq"

\# start MI

set.seed(9202020)

imp <- mice(d1key,m=25)

implist <- imputationList(mids2datlist(imp))

\# construct stratification variables from imputed values

implist2 <- within( implist, {

strata1 = factor(paste(age,race,gender,zip,sep="-"))

strata2 = factor(paste(income,zip,sep="-"))

covid19 = as.numeric(covid=="R")

} )

\# remove imputed strata with no match in population file

implist3 <- subset_datlist( implist2, expr_subset=expression(

strata1 %in% strata1marg$strata & strata2 %in% strata2marg$strata2),

toclass="imputationList"

)

\# Post-stratify on both marginals using raking

design1mi = svydesign(id=~1, data=implist3)

designrakemi = design1mi

\# edit rake function here to make partial=TRUE default, save as rake2

designrakemi$designs <- lapply(design1mi$designs, rake2, sample=list(~strata1,~strata2),

population=list(strata1marg,strata2marg),

control=list(maxit=50, epsilon=.01))

\# Weighted MI

prevs = with(designrakemi, svymean(~covid19))

summary(MIcombine(prevs), digits=3)

\#	results	se (lower upper) missInfo

#covid19	0.133 0.00665 0.119 0.146	72 %

\# Unweighted MI

prevs.nowts = with(implist2, glm(covid19~1))

summary(MIcombine(prevs.nowts), digits=3)

\#	results	se (lower upper) missInfo

\# (Intercept)	0.144 0.00741 0.129 0.159	77 %

set.seed(9202020)

\# Bayesian adjustment for test accuracy

\# U(0,1) prior

parms <- c('sens','spec','prev','testprev')

jagsdata <- list("k" = 351, "Neff" = 2979)	# from d1 unweighted, 11.8%

\# manual initial values, to ensure dispersion

jagsinits <- list(

list("prev" = 0.5, "sens" = 94/101, "spec" = 89/90),

list("prev" = 0.1, "sens" = qbeta(0.1,94,7), "spec" = qbeta(0.1,89,1)),

list("prev" = 0.9, "sens" = qbeta(0.9,94,7), "spec" = qbeta(0.9,89,1))

)	# means or medians, 10th percentile, and 90th percentile of priors

jags.2 <- jags.model('covid-accuracy2.jags.txt',

data = jagsdata,

inits = jagsinits,

n.chains=3,

n.adapt = 5000)

samps2 <- coda.samples(jags.2, parms, n.iter = 10000)

gelman.diag(samps2)

summary(samps2)

\# weighted

jagsdata <- list("p.hat" = 0.1147880, "se" = 0.0048243)	# from m0

jags.4 <- jags.model('covid-accuracy3.jags.txt',

data = jagsdata,

inits = jagsinits,

n.chains=3,

n.adapt = 5000)

samps4 <- coda.samples(jags.4, parms, n.iter = 10000)

gelman.diag(samps4)

summary(samps4)

\# MI with weights

jagsdata.mi <- list("p.hat" = 0.133, "se" = 0.00665)

jags.3 <- jags.model('covid-accuracy3.jags.txt',

data = jagsdata.mi,

inits = jagsinits,

n.chains=3,

n.adapt = 5000)

samps3 <- coda.samples(jags.3, parms, n.iter = 10000)

gelman.diag(samps3)

summary(samps3)

\# prior sensitivity analysis

parms <- c('alpha','beta','prev','testprev')

jagsdata <- list("k" = 351, "Neff" = 2979)	# from d1 unweighted, 11.8%

jags.1 <- jags.model('covid-accuracy1.jags.txt',

data = jagsdata,

n.chains=3,

n.adapt = 5000)

samps1 <- coda.samples(jags.1, parms, n.iter = 10000)

gelman.diag(samps1)

\# Multivariate psrf: 1

summary(samps1)

**eTable 3:** Comparison of social, demographic and survey characteristics among adults who consented to the blood sample and those who consented and provided a blood sample, Orange County, CA, July 10^th^ to August 16^th^, 2020.

| **Characteristic** | **Consented to, but did not provide, blood sample** | | **Consented to, and provided, blood sample*** | |
| --- | --- | --- | --- | --- |
|  | **N** | **(%)** | **N** | **(%)** |
| **Age** |  |  |  |  |
| 18 to 34 years | 503 | (31.4) | 677 | (22.6) |
| 35 to 54 years | 707 | (44.2) | 1,367 | (45.7) |
| ≥55 years | 391 | (24.4) | 947 | (31.7) |
| **Gender** |  |  |  |  |
| Female | 904 | (56.6) | 1,685 | (56.4) |
| Male | 694 | (43.4) | 1,303 | (43.6) |
| **Race/Ethnicity** |  |  |  |  |
| Hispanic (including multi-race) | 881 | (55.0) | 1,115 | (37.3) |
| Asian (Non-Hispanic, Non-African American) | 205 | (12.8) | 471 | (15.7) |
| Other Non-Hispanic (including white and African American) | 515 | (32.2) | 1,405 | (47.0) |
| **Household Income per year** |  |  |  |  |
| <$50,000 | 595 | (37.2) | 656 | (21.9) |
| $50,000 to $99,999 | 357 | (22.3) | 725 | (24.2) |
| ≥$100,000 | 308 | (19.2) | 1,095 | (36.6) |
| Prefer not to answer | 341 | (21.3) | 515 | (17.2) |
| **Had contact with anyone suspected or confirmed of COVID-19?** |  |  |  |  |
| Yes | 335 | (21.0) | 581 | (19.4) |
| No | 1,085 | (67.9) | 1,857 | (62.2) |
| Don’t know | 178 | (11.1) | 549 | (18.4) |
| **Had any SARS-CoV-2 symptoms in last 2 months?** |  |  |  |  |
| Yes | 495 | (34.8) | 1,040 | (34.8) |
| No | 1,106 | (65.2) | 1,951 | (65.2) |

* Denominators in this column are slightly larger (0.4%) than those provided in Table 1 of the main manuscript owing to the fact that CoVAM results were conclusive for 99.6% (but not all) of participants who provided a blood sample.

**Incidence Fatality Risk**

Estimation of SARS-CoV-2 Incidence Fatality Risk among Adults in Orange County, July 10 to August 16, 2020

We estimated SARS-CoV-2 incidence fatality risk among adults, by age, in three ways.

1. **Baseline**

First, we used a "baseline" estimate which counts the number of SARS-CoV-2 deaths and cases directly from OCHCA's publicly available dataset (https://occovid19.ochealthinfo.com/coronavirus-in-oc). This estimate does not require any population-based seroprevalence estimates from our study. We used August 16th as the last date of counting both cases and deaths, since August 16th is the last date in which we retrieved blood specimens for the seroprevalence test. We categorized cases and deaths into two age groups: 18 to 64 years and 65 years and above.

Deaths (OCHCA Report)
 18 to 64: 224
 65+:        586

Adult Cases, as of 8/16/20 (n=42,953, OCHCA Report)
Estimated cases, by age group:
 18 to 64: 37,369
  65+:       5,584

OCHCA incidence fatality risk, 8/16 (OCHCA Report-based numerators and denominators)
18 to 64: **0.00599**  (i.e., 224 / 37,369)
 65+:       **0.105**      (i.e., 586/ 5,584)

2. **Seroprevalence Estimates and Reported SARS-CoV-2 Deaths**

**Second,** we used the same death counts from the OCHCA dashboard but applied as the denominator the estimated total population infected with SARS-CoV-2 from our seroprevalence survey. Prevalence was relatively consistent across the two age groups, at 11.5 to 11.8 percent. We applied the age-specific prevalence to the estimated population counts for ages 18-64 and 65 and over, from the July 1, 2019 US Census Estimates for Orange County (https://www.census.gov/quickfacts/orangecountycalifornia).

Total Adult Population in OC

18 to 64: 2,000,686
 65+:          485,881

SARS-CoV-2 prevalence based on UCI survey
18 to 65:  11.83%  (302 / 2553)
65+:         11.50%     (49 / 426)

Estimated total population infected with SARS-CoV-2:
 # 18 to 64:  236,681
 # 65+:          55,876

Population-based incidence fatality risk, 8/16 (OCHCA-based numerators and population-based survey denominators)
18 to 64:  **0.00095**  (i.e., 224 / 236,681)
 65+:        **0.0105**   (i.e., 586 /  55,876

3. **Seroprevalence Estimates and Excess Deaths**

Third, we used excess death counts, by age group, in OC under the assumption that cause-of-death data on SARS-CoV-2 underestimate the deaths attributable to SARS-CoV-2.  Use of excess death counts may capture mortality that is above levels expected from seasonality and trend and which likely arises from complications due to SARS-CoV-2. Demographers recommend the use of excess mortality when deriving plausible ranges of incidence fatality rates.^8^ We, as in the Second approach, used as population denominators the estimated total population infected with SARS-CoV-2 from our seroprevalence survey.

 Deaths (Excess)
18 to 64: 441.8
 65+: 650.6

Population-based incidence fatality rate using Excess Deaths 8/16 (Excess)
18 to 64: **0.00187**  (i.e., 441.8 / 236,681)
 65+:       **0.0116**   (i.e., 650.6 /  55,876)

SUMMARY

Calculations from the three different approaches gives a range of infection fatality risks that are:

 for 18 to 64 year olds: **0.95 to 1.9 deaths per 1,000 persons**

 for 65 years and above: **10.5 to 105 deaths per 1,000 persons**

Put another way, for 18 to 64 year olds:

- the SARS-CoV-2 infection fatality risk using seroprevalence estimates is **3.2 to 6.3 fold lower** than that calculated from reported OCHCA statistics.

Also, for 65 years and above:

- the SARS-CoV-2 infection fatality risk using seroprevalence estimates is **9.0 to 10.0 fold lower** than that reported by OCHCA statistics.

An important caveat involves the fact that CoVAM detects infection with doses of SARS-CoV-2 that may fall well below levels detected from other assays. We also, as with other tests, do not know whether and to what extent exposure to SARS-CoV-2 confers immunoprotection against re-infection.
