## Supplementary figures and images for "Estimated Seroprevalence of SARS-CoV-2 Antibodies Among Adults in Orange County, California"

### Supplemental_Material_CoVAM_map

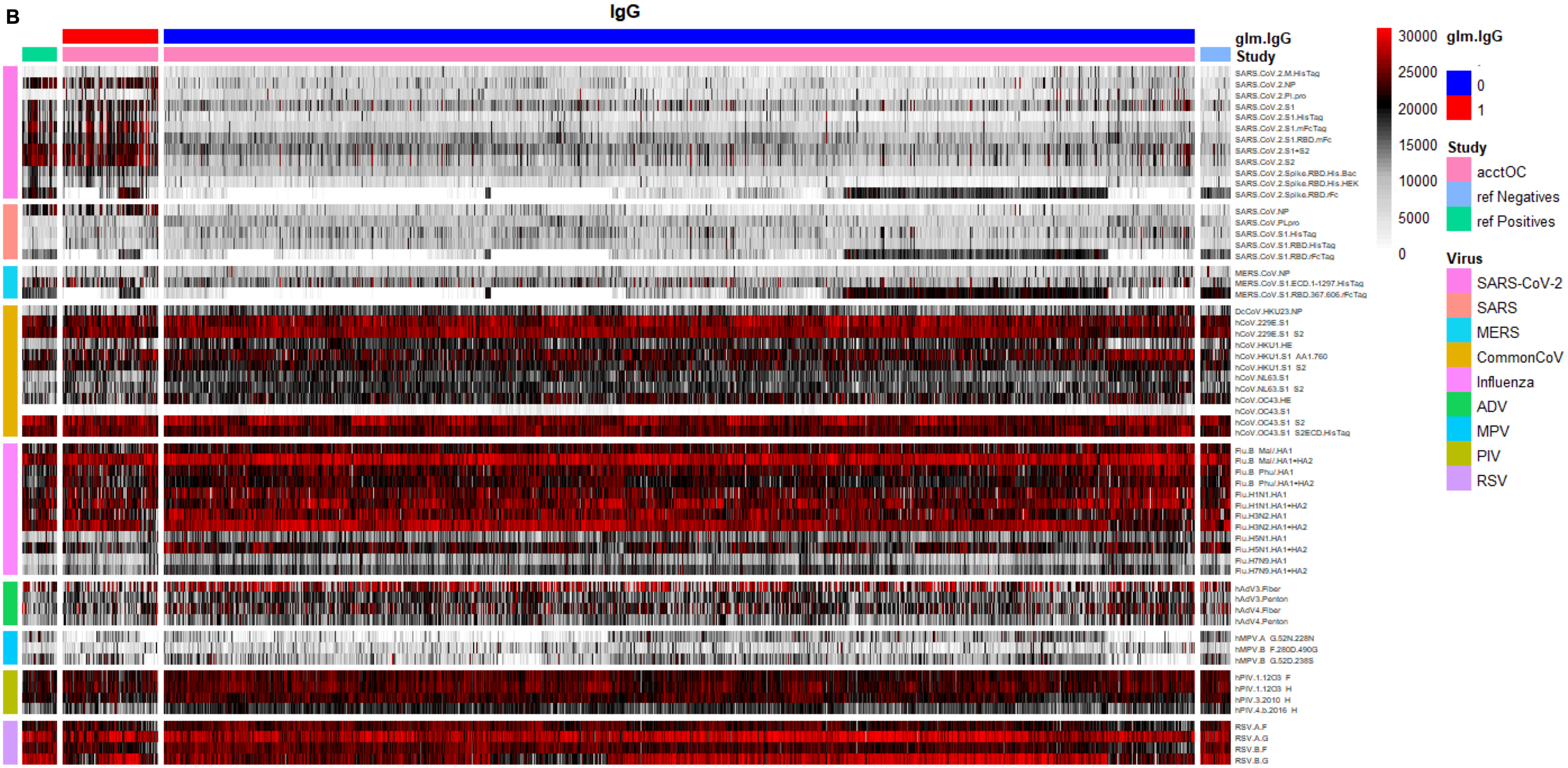
